# Risk factors at birth, in childhood and adolescence for the onset of musculoskeletal chronic pain in adulthood: a scoping review

**DOI:** 10.1101/2021.04.29.21256313

**Authors:** Margelli Michele, Alice Gianelli, Lucia Borghi, Federico Zanchetta, Stefano Salvioli, Maselli Filippo

## Abstract

**Background:** Literature already confirmed that chronic pain is not only correlated to functional deficits, structural problems or neural alterations, but it is also strongly influenced by the presence of psychological and social factors. It was also observed that those factors can have an impact on the subject even after many years: in fact, pain experiences and traumatic events during childhood can lead to changes in stress and pain neural network area and to maladaptive behaviors that concur to develop chronic pain. The main purpose of this review is to identify the bio-psycho-social risk factors in developmental age that could lead to musculoskeletal chronic pain in adulthood.

**Inclusion criteria:** Every study about risk factors in developmental age (0-18) for the onset of musculoskeletal chronic pain in adulthood will be included. This scoping review will consider studies conducted in any context.

**Methods:** The proposed scoping review will be conducted in accordance with the Joanna Briggs Institute methodology (JBI) for scoping reviews.

The search will be carried out on 6 databases: *MEDLINE, Cochrane Central, Scopus, CINHAL, Embase, and PEDro*.

Selection and data extraction will be conducted by two blind independent researchers and inconsistencies will be resolved by a third reviewer.

The results will be presented in a schematic, tabular and descriptive format that will line up with the objectives and scope of the review.

**Conclusions:** This will be the first scoping review to provide a comprehensive overview of the topic. The results will add meaningful information for clinicians especially during the patient’s assessment. Furthermore, any knowledge gaps of the topic will be identified. The results of this research will be published in a peer-reviewed journal and will be presented at relevant (inter)national scientific events.

## Background

Musculoskeletal chronic pain is a very significant problem not only from an epidemiological, economic and social aspect, but also because it leads to a poorer quality of life^1^.

According to literature, chronic pain is not correlated only to functional deficits, structural problems or neural alterations, but it is strongly influenced by the presence of psychological and social factors^2^; indeed these can be both a consequence of the pathology, a cause or modulating factors^3^. The complex dynamics of pain is also related to social life, sleep disorders, depression, anxiety and catastrophization^4^.

These multiple factors influence the experience of pain, making the assessment of these condition extremely complex and heterogeneous^5^. Searching for a patient’s previous pain experience and family history represents a significant part of the evaluation about this condition^6,7^; indeed literature suggests that during childhood, the individual is subject to greater plasticity in brain development by external stimuli^8^. It has been observed that painful stimuli during the neonatal period can cause long-term neuroplastic changes in stress and pain neural network area^9^. These changes in childhood, lead to maladaptive behaviors that concur to develop chronic pain^10^. Moreover, literature emphasizes the importance of emotional abuse and childhood maltreatment, which can induce longterm changes in adult somatosensory function^11^.

Furthermore, childhood abuse survivors demonstrate greater neuroendocrine stress reactivity, suggesting an increased vulnerability to stress-related illnesses^12^.

This means that adverse childhood events have both a biological and a psychological influence on the subject, encouraging the development of physical disorders but also behavioral disorders. There are already some studies that describe the risk factors in childhood that could develop chronic pain during adulthood^6,7,13,14^, but they are heterogeneous and analyze just a single risk factor at once. Knowing and clarifying risk factors in this topic may have direct impact on pain, on his treatment and on prevention of chronicity. The aim of this scoping review is to chart and describe every bio-psycho-social factor that has been considered in literature to this topic.

### Review question

What are the risk factors present in developmental age, from birth to 18 years, for the onset of chronic musculoskeletal pain in adulthood?

The main objectives of this study will be :

1. To map available literature on genetic, neurobiological, psychological factors, social pain learning factors, parenting style and stressful environment during developmental age^15^ that can promote the onset of musculoskeletal chronic pain in adults.
2. To describe final results according to the type of studies and the type of factors (analyzed individually or as a whole), and the tools or scale used to evaluate them.
3. To propose an overall classification of these risk factors and their psychometric values.
4. To identify any possible knowledge gaps on this topic.

## Methods

### Study design and protocol

The proposed scoping review will be conducted in accordance with the Joanna Briggs Institute methodology (JBI) for scoping reviews^18^.

The Preferred Reporting Items for Systematic reviews and Meta-Analyses extension for Scoping Reviews (PRISMA-ScR) Checklist for reporting will be used^19^, and it is priori registered at Medxriv (https://www.medrxiv.org).

### Search strategy

The search will be carried out on 6 databases: *MEDLINE, Cochrane Central, Scopus, CINHAL, Embase, and PEDro*. Studies will also be included searching in the bibliography of relevant revisions, and in Google Scholar. Further research of Gray literature will be carried out through open gray.eu, a multidisciplinary European database. Keywords inserted are: *Childhood, adolescent, infant, young, adult, human, risk factors, chronic pain*. As recommended in all JBI types of reviews and PRISMA-S^20^, a three-step search strategy should be developed:

1. The first step is represented by an initial limited search of an appropriate online database which is relevant to the topic (MEDLINE). This research is based on identifying an appropriate search string by following the acronym PCC (Population, Concept and Context) proposed by The JBI^18^ and using the keywords found by the preliminary background search. The initial search is followed by the analysis of the text words contained in the title, of the retrieved papers’ abstract, and of the index terms, used to describe the articles, on SR accelerator (https://sr-accelerator.com).
2. The second step concerns a search based on all identified keywords and index terms that should be undertaken across all included databases.
3. The third step, the reference list of identified reports and articles should be searched for additional sources.

The search strategies were peer-reviewed by an experienced librarian and were further refined through team discussion. No search limitations and filters applied (language and time). Reviewers’ intent to contact authors of primary sources or reviews for further information. Complete search strategy for Medline is included as an appendix 1 to the protocol. Search strategy will be adapted to be used in other databases.

### Inclusion criteria

We will follow the acronym PCC to describe elements of the inclusion criteria:

#### Population

This scoping review will consider studies with adults (> 18 years old) with musculoskeletal chronic pain, in 1 or more anatomic regions that persists or recurs for longer than 3 months^16^.

#### Concept

This scoping review will consider research studies about risk factors in developmental age (0-18)^17^, for the development of musculoskeletal chronic pain in adulthood: birth, infant (0-2), childhood (early and late) (2-11), adolescent (12-18).

#### Context

This scoping review will consider studies conducted in any context.

#### Sources

This scoping review will consider any study designs or publication type for inclusion. No date and geographical limits will be used. We will consider articles in English, Italian and French; or articles of any language, with at least available English abstract. Studies that do not meet the above-stated Population-Concept-Context (PCC) criteria or which provide insufficient information will be excluded. The rational for choice is part of our initial application.

### Study selection

For the selection process, the first thing to do is to select a sample of 25 title/abstracts that will be analyzed from the entire team using eligibility criteria and definitions/elaboration document. Then, the team will discuss discrepancies and make modifications to the eligibility criteria and definitions/elaboration document. The team will only start screening when 75% (or greater) of agreement between reviewers will be achieved. To start the screening, we will use the Rayyan online software (https://www.rayyan.ai).

The selection phase will begin assessing all the titles and abstracts of the studies retrieved using the search strategy and those from additional sources. These studies will be screened independently by two review authors, to identify those that potentially meet the inclusion criteria. Any disagreement will be solved by discussion with a third author. Subsequently, the full text of these potentially eligible studies will be independently assessed for eligibility by the two reviewers. The reasons for excluding articles will be recorded. We developed a google form (charting table) containing the elements to standardize the selection.

There should be a narrative description of the selection process accompanied by a flowchart of review process (from the PRISMA-ScR statement).

### Data extraction

Extraction module (in appendix B) will be reviewed, before the implementation, by the research team and pre-tested to ensure that the form accurately captures the information. Modifications will be detailed in the full scoping review.

Data extraction will be conducted by two blind independent researchers and inconsistencies will be resolved by a third reviewer.

### Data items

Key information will be described in a charting table with the description of: Author; Publication year; Place of study’s conduct; Setting of study’s conducts; Methodology/ type of study; Aims of the study; Population, Characteristics from which patients are extracted, including gender and age, social variables; Concept: childhood risk factors to develop adult chronic pain; Type of chronic pain; Comorbidity; Psychological factors; Social factors; Outcome measures used for childhood factors

### Critical appraisal of individual sources of evidence

Not provided.

### Data management

As a scoping review, the purpose is to aggregate the findings and to present an overview of the research rather than to evaluate the quality of the individual studies. The results will be presented as a map of data, which are extracted from different documents. These will be included in a schematic, tabular and descriptive format that will line up with the objectives and scope of the review. Descriptive analysis will consist on a distribution of the evidence sources by year or period of publication, countries of origin, area of intervention (clinical, political, educational, etc.) and research methods. Results will be presented such as: population, risk factors and the tools used to assess them, age class. An overall risk factors classification will be proposed with narrative description and table.

## Data Availability

Due to the nature of this research, participants of this study did not agree for their data to be shared publicly, so supporting data is not available.

## Appendix A Complete search strategy

((adult*[Title/Abstract] OR adulthood[Title/Abstract] OR “Adult”[Mesh]) AND (“Chronic Pain”[Mesh] OR “chronic pain”[Title/Abstract] OR “widespread pain”[Title/Abstract] OR “long term pain”[Title/Abstract] OR “musculoskeletal pain”[Title/Abstract])) AND ((hospitalization[Title/Abstract] OR “social learn*”[Title/Abstract] OR “Child Abuse”[Mesh] OR “Adverse Childhood Experiences”[Mesh] OR “Adult Survivors of Child Abuse”[Mesh] OR “Risk Factors”[Mesh] OR “risk factor*”[Title/Abstract] OR “predisposing factor*”[Title/Abstract] OR “adverse experience*”[Title/Abstract] OR trauma*[Title/Abstract] OR “traumatic event*”[Title/Abstract] OR adversit*[Title/Abstract] OR abuse[Title/Abstract] OR parent*[Title/Abstract] OR “genetic factor*”[Title/Abstract] OR “Social Learning”[Mesh] OR “Hospitalization”[Mesh]) AND (“Child”[Mesh] OR “Infant”[Mesh] OR “Adolescent”[Mesh] OR newborn[Title/Abstract] OR childhood[Title/Abstract] OR offspring*[Title/Abstract] OR child[Title/Abstract] OR infant*[Title/Abstract] OR adolescent*[Title/Abstract] OR young[Title/Abstract] OR youth[Title/Abstract] OR children[Title/Abstract] OR birth[Title/Abstract] OR pediatric[Title/Abstract]))

**Appendix B.**
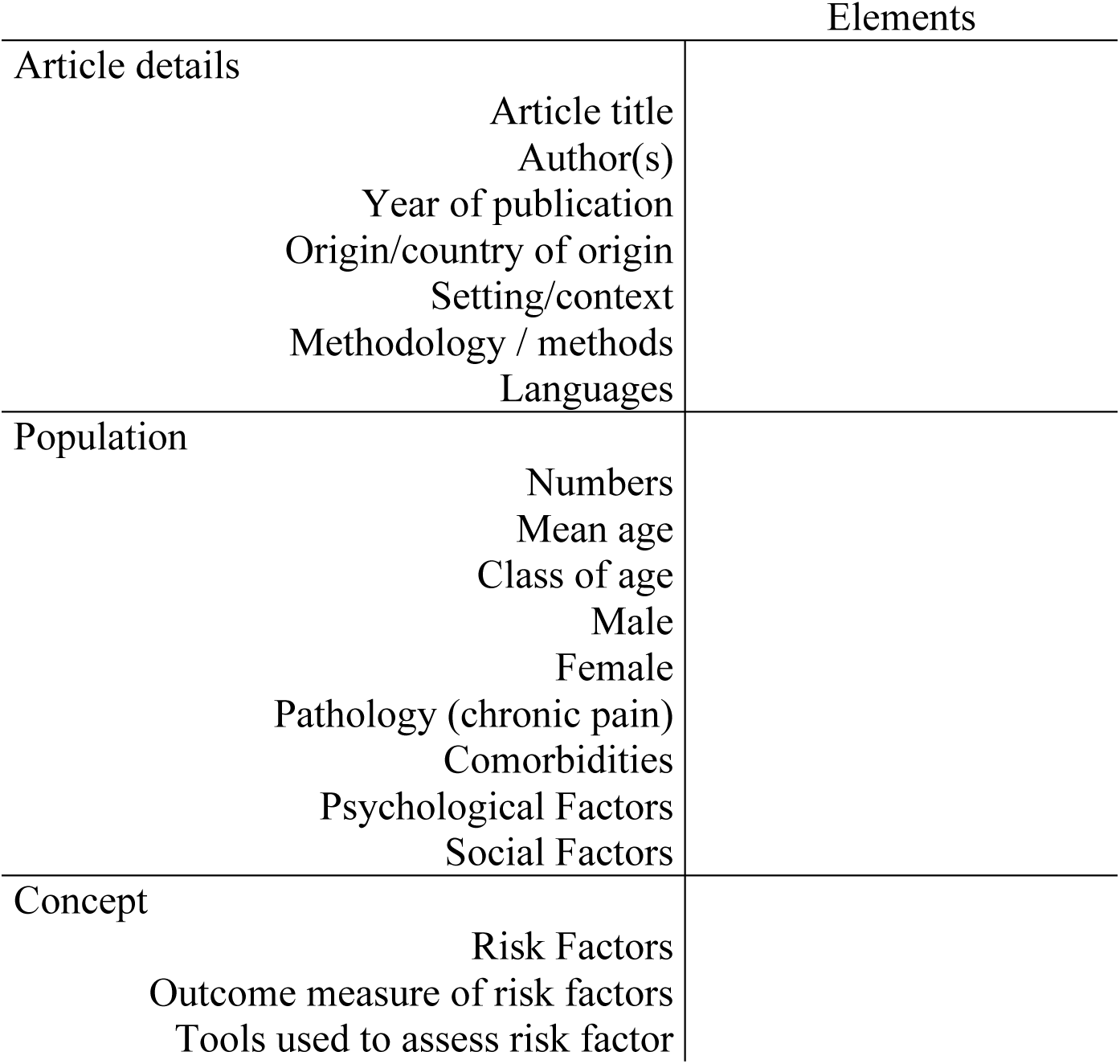
Extraction module.

| Elements |  |
| --- | --- |
| Article details | Article title<br>Author(s)<br>Year of publication<br>Origin/country of origin<br>Setting/context<br>Methodology / methods<br>Languages |
| Population | Numbers<br>Mean age<br>Class of age<br>Male<br>Female<br>Pathology (chronic pain)<br>Comorbidities<br>Psychological Factors<br>Social Factors |
| Concept | Risk Factors<br>Outcome measure of risk factors<br>Tools used to assess risk factor |

